# Two effects of hormone replacement therapy: Explaining the breast cancer risk paradox

**DOI:** 10.1101/2024.02.16.24302914

**Authors:** Kathrin Halfter, Anne Schlesinger-Raab, Dieter Hölzel

## Abstract

**Background:** Hormone replacement therapy (HRT) is currently linked to increased breast cancer (BC) diagnoses. While this appears paradox to an estimated 15-year development period of hormone receptor-positive BC, the discrepancy can be explained if HRT has two effects.

**Methods:** We modelled cohorts of 100,000 women using two parameters: HRT caused accelerated tumour growth and increased incidence. A reference cohort used age-specific incidence, a 15-year growth period, and life expectancy. Treatment cohorts with faster growth and higher incidence were simulated over 30 years. Study endpoints were cumulative and annual BC incidence. Using data from the Womens Health Initiative (WHI), we estimated the factors that reproduce the WHI long-term outcomes.

**Results:** The data indicate that HRT accelerates the growth of previously undetected BC, causing them to appear as seemingly new cases. The timing and duration of HRT determine when this rise occurs. After roughly 15 years of tumour development, an inherent HRT-related risk becomes apparent and overlaps with the expected baseline risk of aging women. The WHI results were reproduced with a growth acceleration factor of 1.4 and an initiation risk factor of 2, consistent with an approximate 10% drop in BC incidence in the United States around 2002 following reduced HRT use.

**Conclusion:** Estimates of about one million additional HRT-associated BCs may largely reflect accelerated growth of pre-existing BCs. Risk-adapted screening strategies could diagnosis newly initiated tumours early. By communicating these short- and long-term effects of HRT on cancer risk patients and physicians could make an informed decision on the risks and benefits of HRT-use.

## Introduction

Hormone replacement therapies (HRT) have been used for over 70 years to alleviate menopausal symptoms.^1,2^ Despite their efficacy, the risk-benefit discussion remains highly controversial. This debate is fueled by reports of increased risks for breast and endometrial cancer, dementia, and stroke. However, data also confirm protective effects, such as a reduced risk of diabetes, bone fractures, and colorectal cancer.^3^ Consequently, HRT remains a central, medically indicated treatment for severe menopausal symptoms.^4^

A critical turning point occurred in 2002 when the Women’s Health Initiative Study (WHI-Study) reported an increased breast cancer (BC) risk associated with combined HRT (estrogen and progesterone).^5^ This publication led to a sharp decrease in HRT usage and a decline in BC incidence in the USA.^5,6^ Further crucial data linking HRT to BC risk emerged from the Million Women Study (MWS)^7^ and comprehensive Meta-Analyses (Meta-A).^8,9^ Meta-A estimated that approximately 1 million BC cases in Western countries since 1990 were attributable to HRT use within the first five years of therapy.^9^

While numerous studies have investigated BC risk, analyses of the underlying biological processes are lacking. Since hormone receptor-positive (HR+) BC takes approximately 15 years to become detectable, the observed increase cannot be explained by short-term BC promoting factors. The connection can, however, be understood via two distinct mechanisms: growth acceleration of pre-existing BC lesions and initiation of *de novo* BCs. The aim of this model study was to quantify these two effects and their interactions. The results aim to enable a more nuanced communication of the short- and long-term health effects of HRT, supporting risk management through adjusted screening.

## Methods

Four variables are required for the simulation of HRT-related tumor growth and BC risk:

* Tumor growth
* Growth acceleration factor (GF) = 2
* HRT duration = 4 years
* Tumor-initiating risk factor (RF) = 2

The BC volume doubling time (VDT) depends on HR status: 72 days in HR– and 170 days in HR+ tumors. Considering the subtype distribution (HR+ 85% /HR– 15%), a weighted mean VDT of 150 days is plausible.^10–12^ Growth duration from diameter TD1 to TD2 for HR+ tumors is based on the equation:

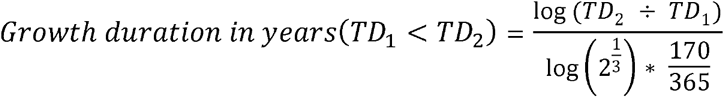

(TD tumor diameter in mm, 1 tumor cell ≈ 0.01mm or 10 µm. Growth to 20 mm takes ≈ 15.3 years (≈33 VDs)). The interval from 5–20 mm, where most screen-detected cancers lie, requires ≈ 2.8 years.

The chosen parameters are plausible approximations based on published studies (see supplement table S5 for data sources and underlying assumptions). GF=2 is smaller than the HR+/-ratio 170/72 and RF=2 is based on the Gail model and was used for recruitment in chemoprevention trials.^13,14^

Four virtual cohorts are generated:

### 1. Control cohort (n=100,000, female age 50)

Based on the SEER incidence from 2000 and life expectancy, BCs were randomly generated for 30 years up to the age of 80. Because studies require a negative mammogram at baseline, only half of the ≈250 expected BCs are randomly assigned in the second half of the first year. Over the observation period, 9,961 BC cases and 27,612 non-cancer deaths occur; total person-years without BC survival equals ca. 2.67 million. The 15-year growth period allows estimation of tumor onset before or after HRT initiation.

### 2. Cohort: HRT with growth acceleration

The control cohort undergoes HRT for 4 years. Prevalent tumors grow twice as fast (GF=2), superimposed on the development seen in cohort 1. An additional assumption is that HRT acts like an on/off switch.

### 3. Cohort

HRT with tumor initiation + growth acceleration: RF=2 is added to simulate *de novo* BC initiation. Both prevalent and newly initiated tumors experience accelerated growth during years 0–4 of HRT.

### 4. Cohort

Endocrine prevention cohort: This arm simulates the BC risk reducing effect of endocrine prevention on control cohort 1 for comparisons with the HRT risk.^15^. Cohort 3 reflects the combined effects of RF and GF. The difference in cumulative incidence from cohort 1 yields the RR. The RR due to GF appears during HRT as a short-term incidence increase, while the hazard ratio (RR) due to RF becomes detectable only ≈20 years later due to tumor growth time. This generation process was used to simulate the WHI-Study. Age distribution, sample size, incidence in USA during the 2000s, and RR were specified; GF and RF were iteratively adjusted until simulated cumulative incidences matched the reported RRs. Results were compared with published data. The modelling approach follows recommended standards.^16^ Statistical analyses were performed using R version 3.1.3.

## Results

Fig. 1. shows the timeline when BCs were initiated and detected with or without HRT. Three periods can be distinguished in which the diagnosis of BCs is shifted forward by HRT. The oldest prevalent BCs, which would normally be diagnosed within the first 4 years, are advanced by up to 2 years under HRT with a GF of 2. In Period 2, all BCs occur two years earlier, resulting in no change in incidence compared with cohort 1. Period 3 is prolonged by HRT: the first BC initiated under HRT appears 2 years earlier after 13 years, while the final BC remains unaffected and occurs after 19 years. With a RF of 2, incidence in Period 3 doubles. In Period 4, no changes occur.

**Figure 1.**
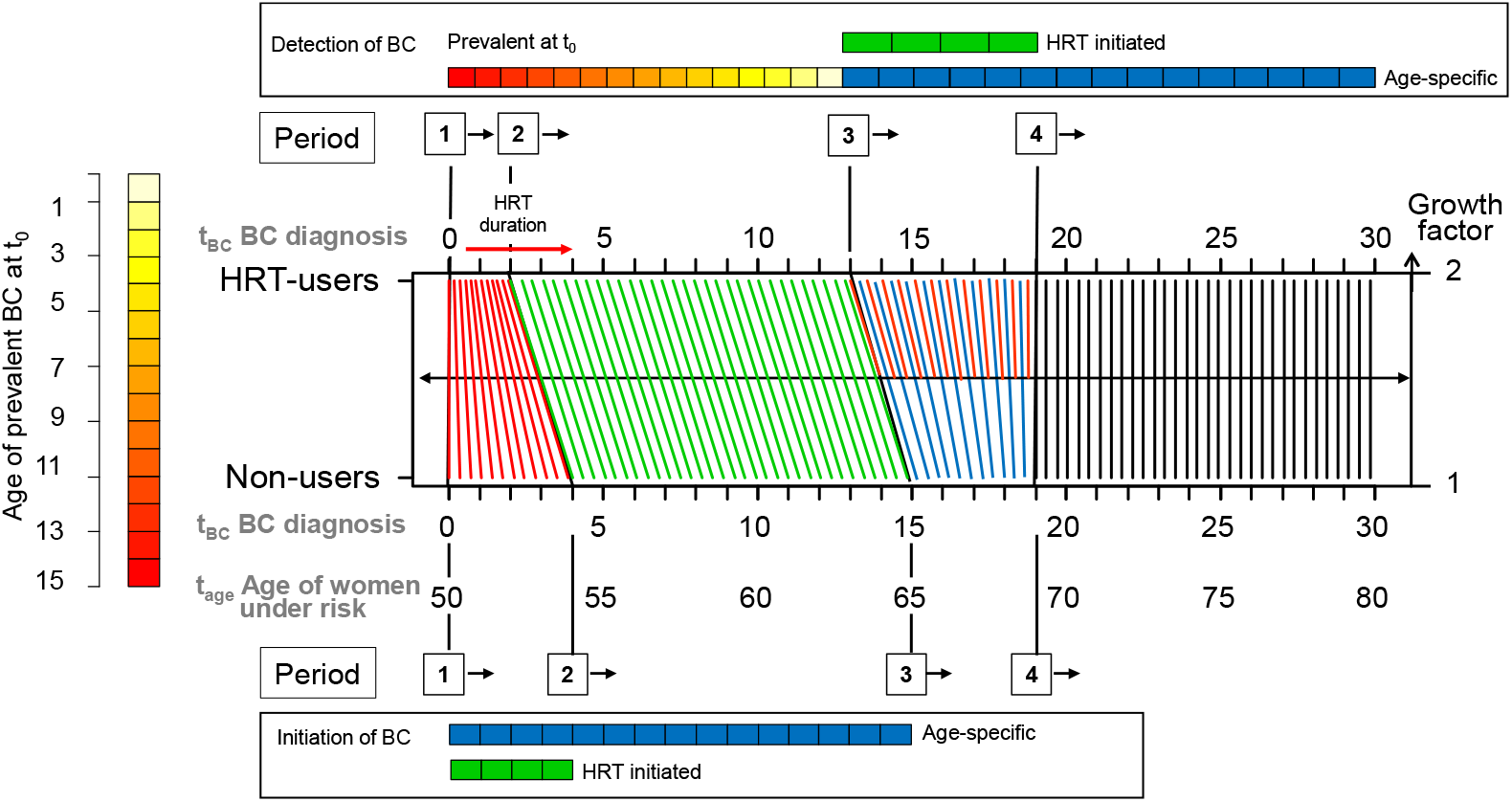
Tumor growth and growth acceleration by HRT. With growth duration of BCs from initiation to detection of 15 yrs, HRT duration of four years, HRT caused growth acceleration with factor 2 and breast cancer risk factor. The effect of HRT (top) is shown in comparison to Non-HRT users (bottom). Transformation of the time of diagnosis for BC by HRT over three decades: Four periods can be distinguished: Period 1 – The growth accelerating effect of HRT leads to an earlier detection of prevalent tumors and earlier metastasis over a time of *HRT intake duration divided by growth acceleration factor*. This leads to an increased BC incidence over this period (two years). For Non-HRT users, the general age-specific BC-risk of 50-54 years old women works over the whole period of HRT duration in the HRT-user group (here: four years). Period 2 – The general age-specific BC risk works in both groups until newly initiated BCs can be diagnosed, this period lasts over the time of *growth duration of BCs minus HRT duration multiplied by ((growth acceleration factor-1) divided by growth acceleration factor)*. This period is shifted forward in HRT-users by *HRT duration divided by growth acceleration factor* (2⍰13 years). In HRT-users the incidence decreases because tumor detection was brought forward due to accelerated growth (green lines). Period 3 – HRT-initiated new BCs (red lines) are diagnosed in addition to forward shifted age-specific BC in HRT-users, the incidence increases. In Non-HRT-users the general age-specific risk works alone. Period 4 – The HRT effects have ended. In both groups again only the general BC risk works (black lines). These transformations and relative risks can be easily calculated if an annual incidence of e.g. 100 is assumed.

Figures 2A–B show the changes in cumulative incidence over 30 years for the four cohorts and provide a more detailed view of the first seven years. The BC incidence of control cohort 1 without HRT corresponds to the general age-specific incidence. In cohort 2 under HRT, period 1 shows an increase, with GF 2 producing a doubling of incidence due to the growth acceleration of larger prevalent tumours (red boxes). Period 1 ends after two years. The effect of the accelerated growth of all prevalent BCs after ≈ 15 years is almost entirely compensated by the BC risk.

**Figure 2.**
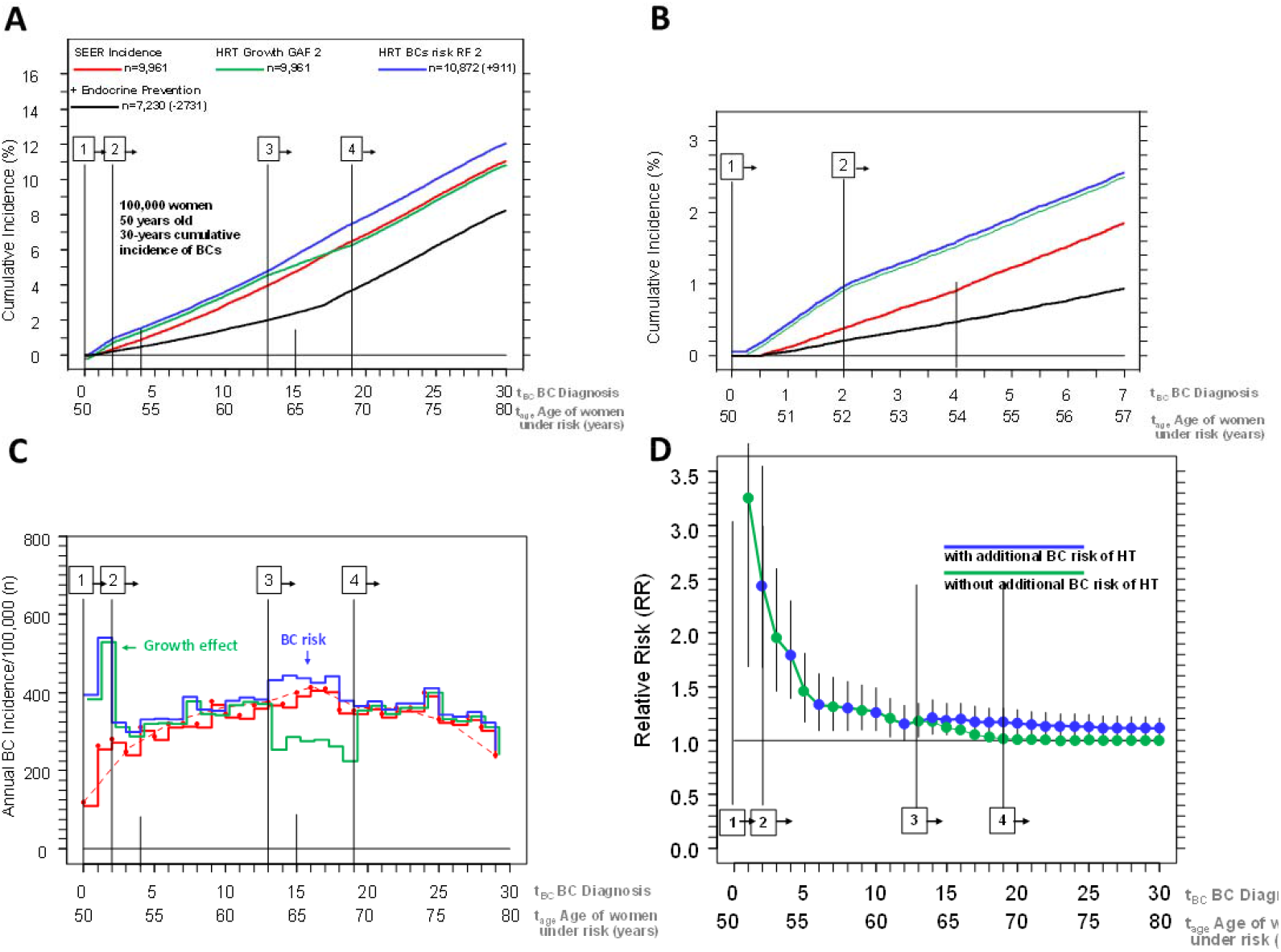
Simulation of incidence for 100 000 women aged 50 years with the model parameters growth duration of BCs 15 years, HRT duration 4 years, growth acceleration factor (GAF) and breast cancer risk factor (RF) of 2 each and life expectancy. (Overlapping lines were shifted manually) **A:** Cumulative incidences generated according to the SEER data from 2000 (red) with 9961 BCs or 11% BCs over 30 years, with faster growth during four years of HRT (green) with the same number and with an additional breast cancer risk (n=911) due to HRT (blue) and with two years of endocrine prevention and 50% BC reduction, which could avoid 2731 or 27.4% of prevalent BCs in 17 years (black). The four periods according to Figure 1 re marked. **B:** The first 7 years of cumulative incidences of Figure 2A enlarged. Due to the inclusion criterion “negative mammography”, almost no BCs are diagnosed in the first six months without HRT, but the first faster-growing BCs are diagnosed after three months with HRT. **C:** Contours of the bar charts of the annual BC number: without HRT (red), with HRT and GAF 2 (green), with HRT, GAF 2 and breast cancer RF 2 (blue). The beginning and end of HRT leads to four periods: At the beginning by growth acceleration of all prevalent BCs, shifting their diagnosis forward by 2 years, and with a time delay for the BCs occurring age-specifically under HRT and those initiated by HRT, both of which are accelerated during HRT and occur or are diagnosed in Period 3 over about six years. The annual fluctuations are due to random generation (Poisson distributed: Standard deviation ±19). The mammography effect with the exclusion of detectable prevalent BCs from the cohorts is evident in the first year. The absolute number of annual BCs decreases with age despite increasing incidence because 27 612 women do not reach the age of 80 due to competing risks. **D:** Time dependence of the complete relative risks with (cohort 3, blue) and without additional breast cancer risk (cohort 2, green) due to HRT and 95% confidence intervals. The risks differ from the 13th year onwards with the additional diagnosis of HRT-initiated new BCs. The inclusion criterion “negative mammography” and growth acceleration factor 2 result in the increased initial risk.

Figure 2C displays the fluctuations in annual incidence rates due to random generation. In cohort 1, incidence rates initially rise according to age-specific patterns and later decline slightly due to reduced numbers of patients at risk as life expectancy decreases. The annual incidence rates in cohorts 2 and 3 illustrate how both modifying factors affect incidence over time.

Figure 2D shows, for each follow-up year, the RR of cohorts 2 and 3 compared with the control group. RR decreases from the 2nd to the 12th follow-up year because cumulative incidence increases at the same annual rate in all three cohorts. Afterward, the RR of cohort 3 rises again due to the HRT-associated BC risk. In cohort 4, two years of endocrine prevention are assumed and simulated. Under the assumption of 50% treatment efficacy, an estimated 2,133 of 4,278 prevalent BCs developing over 15 years would be prevented.

To model the randomized WHI study the following parameters were specified^10^: A total of 8,506 women were recruited for combination HRT. Discontinuation of the study after 5.6 years was taken as HRT duration. BCs in the control group were generated using data on age distribution, life expectancy, and the inclusion criterion of a negative mammogram. The cumulative hazard rates after 5.6/24 years were 1.26/1.28.^5,6,17^ The values of the factors were iteratively adjusted until the target RR values were achieved: With a growth acceleration factor of 1.4 and a BC risk factor of 2.0, the WHI study could be replicated (Figure 3).

**Figure 3.**
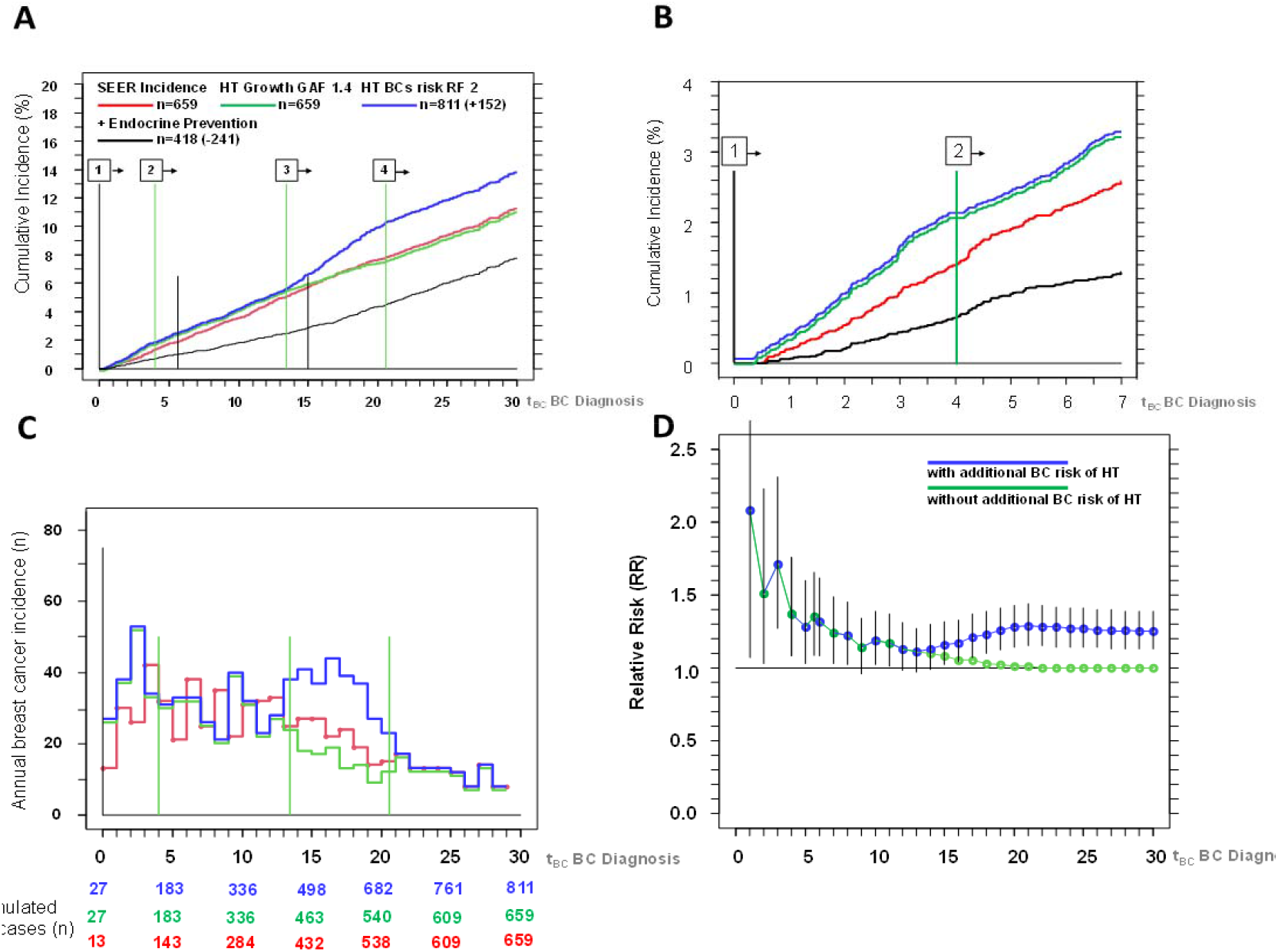
Simulation of the randomized WHI-Study. The model parameters are: 8506 women (n=659/659/811/418 BCs in the four cohorts, see Figure 2), age distribution according to the WHI-Study (MW: 63.6 years), growth duration of BCs 15 years (3313 total years of tumour growth), HRT duration 5.6 years (accelerated tumours n=379, total years of earlier detection = −682 years), negative mammogram, incidence according to SEER data from the year 2000, life expectancy considered as competing risk. They result in the estimates GAF 1.4 and breast cancer RF 2.0 (+152 additional BCs). For a better overview, overlapping lines were shifted manually. **A:** Cumulative incidences of the four groups. Control group of the WHI-Study (red), and the HRT groups without (green) and with breast cancer risks (blue) show the combined effect over 30 years of follow-up. The turning points are at 4, 13.4 and 20.6 years. **B:** Cumulative incidences of the first 7 years enlarged. The effect of the negative mammogram is again illustrated, whereby no BCs occur in the control group in the first six months. In the control group 202 BCs were generated for 7 years, in the first year only 13 BC after day 186, in the HRT group 249 BC were generated and 13 appear before day 186 due to GAF1.4. **C:** Contours of the bar charts of the annual BCs. Due to the higher age of 63.6 years, the number of patients at risk and thus the annual BCs decrease significantly compared to Figure 2C. **D:** Time dependence of the relative risks with and without additional breast cancer risk by HRT and 95% confidence intervals (see Figure 2)

The simulated age distribution yielded a mean age of 63.5 years (WHI: 63.4 years). In the most recent follow-up, the median age after 24 years was 82.0 years, with 3,182 of 8,506 women in the HRT group still at risk. After 8 years, 296/262 BCs (RR 1.22, CI: 1.02–1.45) and after 24 years 761/609 BCs (RR 1.27, CI: 1.14–1.41) were generated in the groups with and without HRT, respectively. These figures compare well with WHI data showing 277/207 BCs after 8 years and 584/447 BCs after 24 years.

The most pronounced discrepancy between the original and replicated studies occurred during the first four years (Figure 3B), when the randomized WHI study even suggested a protective effect of HRT.^6^ However, growth acceleration due to HRT should increase incidence within the first months. This is supported by the WHI observational study of 16,121 women, which reported a hazard rate of 1.55 after 11.3 years and an overall incidence pattern consistent with the simulation (Figure 2).^10^ In the modelled study, 13/27 BCs occurred in the first year (Figure 3C), illustrating the time dependence of the RR with wide confidence intervals due to small case numbers (Figure 3D). The follow-up of 30 years explains the higher absolute number of cases.

The updated Meta-A summarizes data from 58 prospective and retrospective studies, including 143,887 BCs and 424,972 controls.^8,18^ Data from the MWS on BC risk by histological type showed a relative risk of 1.71 for ductal BC after 3.4 years of combination HRT.^7^ Meta-A results demonstrate increasing relative risks of 1.6, 2.26, and 2.51 for HRT durations of 1–4, 10–14, and ≥15 years, respectively, consistent with both modelled effects. In contrast, no simulation showed increased risk after longer HRT exposure. Notably, an apparent reduction in RR to 1.2–1.3 was observed in women during the first five years after cessation of long-term HRT. These findings align with Figure 3D and likely reflect the early phase before the subsequent increase in risk from HRT-initiated BC becomes evident.

## Discussion

HR-positive BCs grow for approximately 15 years before detection (VDT ≈170 days, ≈33 doublings). Chemoprevention studies show that 5 years of endocrine CP reduces the incidence of BCs by almost 20 years. 15 years of growth is also linked to the atomic bombings, because after that, the frequency of BCs among survivors increased. Finally, the tumor-specific survival rate of 95% after 20 years for pT1b BCs indicates that MET of HR+ BCs grow for approximately 10 years and that BCs take longer. On the other side the increase in incidence in the first few months under HRT with estrogen and progesterone is also a fact. However, the assumption that one million de novo BCs were “largely caused” by HRT in the first five years in Western countries since 1990 is paradoxical.^8^ There are no oncological anomalies or deviations from classic tumor characteristics; even triple-negative tumors grow for about six years. There is also no evidence of an increased incidence of invasive prevalent tumors. Autopsy studies show prevalence above expectations only for DCIS, not for invasive cancer. ^19,20^

The assumption of two effects, a growth acceleration (GF) and moderate risk increase (RF), is therefore plausible and suitable for simulating the WHI study results. A GF of ≈1.4 corresponds to observed differences in metastasis growth of HR-positive tumors between KI67 values of 20% and 50%. HRT increases proliferation, KI67, and mammographic density even in healthy breast tissue.^21,22^ An RF of ≈2 is consistent with chemoprevention studies using the Gail model (without BRCA markers).

Growth acceleration also explains the sharp decline in US breast cancer incidence after 2002 following the WHI publication. The rapid decrease suggests that an immediate factor affecting growth acceleration was switched on and off. Estimated incidence fell from 203,500 cases in 2002 to 182,460 in 2008.^23,24^ Among women aged 50–70 years (incidence 356/100,000), a GF of 2 could explain a reduction of ≈21,000 cases (10.3%), consistent with 5–6 million women discontinuing HRT out of all 43.6 – 48.4 million women aged 45-74 in 2002.^25–27^ A similar decline in HRT prescriptions has also been shown for the Welsh population. ^28^ Alternative oncological anomalies, such as an anti-estrogenic effect forcing nearly detectable tumors into permanent dormancy or apoptosis, are biologically implausible and has not been experimentally proven.

This model also accounts for long-term US incidence trends: the temporary increase from 1980s screening continued with rising HRT use until 2000. By 2006, incidence returned to 1986 levels.^29^ This suggests that the peak between 1986 and 2000 largely reflected earlier detection of prevalent tumors. Since growth acceleration does not generate de novo BCs, incidence subsequently returns to baseline. Current SEER data show rates of 132.6/100,000 (2000), 122 (2004), and 126.4 (2014).^30^

The next anomaly is the increased mortality associated with HRT, seen in the MWS but not WHI, due to sample size. Even a doubling of the age-specific incidence does not increase mortality. The accelerated growth, in turn, explains this mortality effect. Accelerated growth leads to larger tumors with more positive lymph nodes and earlier initiation of metastasis. Tumor diameters of 17 and 15 mm were found with and without HRT, respectively ^10^, resulting in relative survival rates of 84.6% and 87.4% after 15 years ^31^. Reliable conclusions require a long follow-up period: HRT-initiated BCs appear after 15 years, and their survival can be assessed after 25 years.^6,10^ Annual screening can offset the lower prognosis among HRT users.^18,32^

A wash-out period must also be considered when interpreting and estimating the time-dependent BC risk. Growth acceleration or tumor initiation due to HRT are not reversible (Suppl Fig.S1); thus, a “safe” wash-out time under 15 years does not exist.^17,33^ HRT use before or during the study must be accounted for in any risk assessment. Likewise, other hormone use (contraceptives, fertility treatment) prior to age 50 should also be considered.

Virtual “twin” studies with the same age distribution and long-term results can be generated for WHI cohorts using population data on HRT use. However, questions remain: Are there biomarkers for HRT-induced tumors? Can temporary growth acceleration be histologically confirmed? Can HRT discontinuation halt or eradicate occult tumors?^2^ What are the effects of different HRT formulations on BC risk, combined estrogen and progesterone versus progesterone-only?

The 2002 incidence decline indicates that millions of women have stopped or avoided HRT due to fear of BCs. Many of them suffering from menopausal symptoms have accepted a reduced quality of life.^3,34^ At five years of HRT, about two prevalent BCs are growth accelerated for every 100 HRT users. As already mentioned, an annual screening for HRT-users could be beneficial. An RF of 2 would also generate two additional cancers per 100 HRT users, expected 14–20 years after starting HRT at age 50. Early detection through screening mammography improves prognosis for these HRT-related tumors.

## Conclusion

The decline in the US breast cancer incidence after 2002 can be explained by discontinuation or avoidance of HRT. The estimated one million HRT-associated cases were not de novo tumors but prevalent BCs that progressed faster and were diagnosed earlier due to HRT. The small additional risk should be weighed against the quality-of-life benefits of HRT in peri- and postmenopausal women. Risk-adapted screening strategies should be offered to HRT users to ensure early diagnosis. The benefits and risks of HRT should be transparent and communicated clearly to allow a shared decision-making process for patients and their treating physicians.

## Data Availability

All data produced are available online at:
http://seer.cancer.gov/
https://seer.cancer.gov/statistics-network/
http://www.tumorregister-muenchen.de/en/facts/specific_analysis.php

https://seer.cancer.gov/statistics-network/

http://www.tumorregister-muenchen.de/en/facts/specific_analysis.php

## List of abbreviations

BC: Breast Cancer
CI: 95% Confidence interval
DCIS: ductal carcinoma in situ
HR: Hormone receptor status
HRT: hormone replacement therapy
MWS: Million Women Study
RR: Relative risk
SD: Standard deviation
TD: Tumour diameter
VDT: Volume Doubling Time
WHI: Women’s Health Initiative

## Declarations

### Ethics Committee approval

Not applicable, use of anonymized retrospective public data. Consent for publication: Not applicable

### Availability of data and materials

The datasets were derived from sources in the public domain under https://seer.cancer.gov/statistics-network/, and publications (Jemal A, Thomas A, Murray T, Thun M. Cancer statistics, 2002. CA: a cancer journal for clinicians 2002; 52(1): 23-47, Jemal A, Siegel R, Ward E, et al. Cancer statistics, 2008. CA: a cancer journal for clinicians 2008; 58(2): 71-96.). All other data underlying the simulation was obtained from publications as referenced.

## Supplementary data

Supplementary data are available at IJE online

## Authors’ Contributions

Concept and design: Hölzel

Acquisition, analysis, interpretation of data: All authors

Drafting of the manuscript: All authors

Critical revision of the manuscript for intellectual content: Halfter, Schlesinger-Raab

Statistical analysis: All authors

Administration, technical, material support: Halfter

Supervision: Hölzel, Schlesinger-Raab

## Use of AI tools

The authors used AI to improve overall readability and grammer.

## Acknowledgment

We would like to thank all former MCR employees for their contributions to the excellent data quality. We would like to thank Steven Narod, Doris Mayr, Sylvia Heywang-Köbrunner, Chistian Thaler and Michael Lauseker for their valuable scientific and technical advice. We thank the heads of the breast cancer project group of the Comprehensive Cancer Centre of the Ludwig-Maximilians-University and the Technical University of Munich for repeated presentations and enriching discussions of results.

