## Supplementak Material for "Two effects of hormone replacement therapy: Explaining the breast cancer risk paradox"

by

Kathrin Halfter^1^ , Anne Schlesinger-Raab^1^, Dieter Hölzel^1^

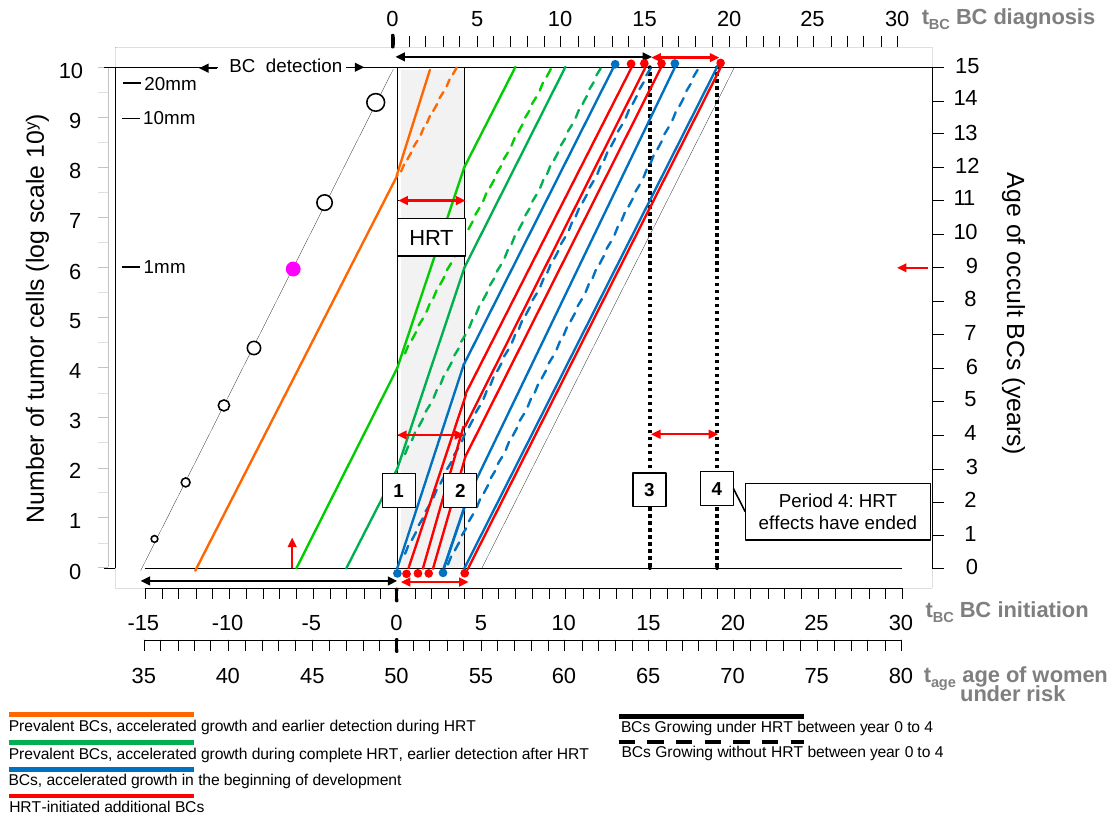

**Figure S1: Examples of BC trajectories**

HRT begins at the age of 50 at time 0. The straight lines represent the number of cells with exponential growth, the right axis shows the “age of development” of the prevalent BCs. If a BC is initiated 15 years earlier, it is excluded due to positive mammography before the start of HRT. The pink dot marks a prevalent 1 mm BC which has grown over 9.3 years from initiation and further six years until detection by mammography, here at time 0. BCs which start between -15 and -11 years (before time 0) grow a maximum of two years faster under HRT (orange trajectory) and are detected during HRT. BCs which are initiated before the start of HRT (green) grow faster over four years of HRT at different tumour ages, their growth slows again after the end of HRT. Their detection is shifted forward in time. The general age-specific BCs developing under HRT (blue trajectories) and additional BCs (red trajectories) developing if there is a breast cancer risk by HRT grow faster under HRT. The first HRT-initiated BCs would appear after about 13 years, the last after 19 years.

**Table S1:** Data on simulated cohort 1: 100 000 50-year-old women under U:S: life expectancy and SEER-age-specific BC risk over 30 years of virtual observation

| **COHORT 1**  **Year of breast cancer diagnosis** | **Women**  **under**  **risk of BC** | **Age-specific**  **new BCs**  **per year ^1^**  **(Cohort 1)** | **Competitive deaths according**  **to life expectancy^2^** | **Expected**  **Survival** |
| --- | --- | --- | --- | --- |
| 0 | 100 000 | 0 | 0 | 1 |
| 0.75 | 99 770 | 63 | 167 | 0.999 |
| 1 | 99 663 | 56 | 51 | 0.998 |
| 2 | 99 145 | 263 | 255 | 0.995 |
| 3 | 98 599 | 281 | 265 | 0.993 |
| 4 | 98 063 | 248 | 288 | 0.990 |
| 5 | 97 426 | 311 | 326 | 0.986 |
| 6 | 96 807 | 287 | 332 | 0.983 |
| 7 | 96 097 | 320 | 390 | 0.979 |
| 8 | 95 335 | 322 | 440 | 0.975 |
| 9 | 94 567 | 319 | 449 | 0.970 |
| 10 | 93 649 | 378 | 540 | 0.964 |
| 11 | 92 771 | 345 | 533 | 0.959 |
| 12 | 91 839 | 342 | 590 | 0.953 |
| 13 | 90 871 | 367 | 601 | 0.947 |
| 14 | 89 886 | 376 | 609 | 0.940 |
| 15 | 88 790 | 370 | 726 | 0.933 |
| 16 | 87 601 | 400 | 789 | 0.924 |
| 17 | 86 348 | 414 | 839 | 0.915 |
| 18 | 85 067 | 410 | 871 | 0.906 |
| 19 | 83 729 | 355 | 983 | 0.896 |
| 20 | 82 306 | 354 | 1069 | 0.884 |
| 21 | 80 777 | 366 | 1163 | 0.872 |
| 22 | 79 229 | 345 | 1203 | 0.859 |
| 23 | 77 621 | 361 | 1247 | 0.845 |
| 24 | 75 867 | 360 | 1394 | 0.830 |
| 25 | 73 966 | 400 | 1501 | 0.813 |
| 26 | 71 999 | 331 | 1636 | 0.795 |
| 27 | 69 900 | 326 | 1773 | 0.776 |
| 28 | 67 657 | 339 | 1904 | 0.755 |
| 29 | 65 187 | 312 | 2158 | 0.730 |
| 30 | 62 427 | 240 | 2520 | 0.702 |
| Total number of BCs / deaths |  | 9961 | 27 612 |  |

**Table S2:** Data on simulated cohort 2: 100 000 50-year-old women under U:S: life expectancy and SEER-age-specific BC risk over 30 years of virtual observation, in addition accelerated growth of prevalent BCs due to HRT (GAF=2)

| **Year of diagnosis** | **Women**  **under**  **risk of BC** | **Age-specific**  **new BCs**  **per year**  **+**  **Growth acceleration due to HRT**  **(Cohort 2)** | **Difference of BC cases compared to cohort 1** | **Competitive deaths according**  **to life expectancy ^1^** | **Expected**  **Survival** |
| --- | --- | --- | --- | --- | --- |
| 0 | 100 000 | 0 | 0 | 0 | 1 |
| 0.75 | 99 586 | 247 | **+184** | 167 | 0.999 |
| 1 | 99 400 | 135 | **+79** | 51 | 0.998 |
| 2 | 98 616 | 529 | **+266** | 255 | 0.995 |
| 3 | 98 040 | 311 | **+30** | 265 | 0.993 |
| 4 | 97 465 | 287 | **+39** | 288 | 0.990 |
| 5 | 96 819 | 320 | **+9** | 326 | 0.986 |
| 6 | 96 165 | 322 | **+35** | 332 | 0.983 |
| 7 | 95 456 | 319 | **-1** | 390 | 0.979 |
| 8 | 94 638 | 378 | **+56** | 440 | 0.975 |
| 9 | 93 844 | 345 | **+26** | 449 | 0.970 |
| 10 | 92 962 | 342 | **-36** | 540 | 0.964 |
| 11 | 92 062 | 367 | **+22** | 533 | 0.959 |
| 12 | 91 096 | 376 | **+34** | 590 | 0.953 |
| 13 | 90 125 | 370 | **+3** | 601 | 0.947 |
| 14 | 89 263 | 253 | **-123** | 609 | 0.940 |
| 15 | 88 252 | 285 | **-85** | 726 | 0.933 |
| 16 | 87 187 | 276 | **-124** | 789 | 0.924 |
| 17 | 86 069 | 279 | **-135** | 839 | 0.915 |
| 18 | 84 936 | 262 | **-148** | 871 | 0.906 |
| 19 | 83 729 | 224 | **-131** | 983 | 0.896 |
| 20 | 82 306 | 354 | 0 | 1069 | 0.884 |
| 21 | 80 777 | 366 | 0 | 1163 | 0.872 |
| 22 | 79 229 | 345 | 0 | 1203 | 0.859 |
| 23 | 77 621 | 361 | 0 | 1247 | 0.845 |
| 24 | 75 867 | 360 | 0 | 1394 | 0.830 |
| 25 | 73 966 | 400 | 0 | 1501 | 0.813 |
| 26 | 71 999 | 331 | 0 | 1636 | 0.795 |
| 27 | 69 900 | 326 | 0 | 1773 | 0.776 |
| 28 | 67 657 | 339 | 0 | 1904 | 0.755 |
| 29 | 65 187 | 312 | 0 | 2158 | 0.730 |
| 30 | 62 427 | 240 | 0 | 2520 | 0.702 |
| Total number of BCs / deaths |  | 9961 | 0 | 27 612 |  |

**Table S3:** Data on simulated cohort 3: 100 000 50-year-old women under U:S: life expectancy and SEER-age-specific BC risk over 30 years of virtual observation, in addition accelerated growth of prevalent BCs due to HRT (GAF=2) and risk of HRT-initiated BCs (RF=2)

| **Year of diagnosis** | **Women**  **under**  **risk of BC** | **Age-specific**  **new BCs**  **per year**  **+**  **Growth acceleration due to HRT**  **+ HRT-initiated new BCs (Cohort 3)** | **Difference of**  **BC cases compared to cohort 1** | **Competitive deaths according**  **to life expectancy ^1^** | **Expected**  **Survival** |
| --- | --- | --- | --- | --- | --- |
| 0 | 100 000 | 0 | 0 | 0 | 1 |
| 0.75 | 99 586 | 247 | **+184** | 167 | 0.999 |
| 1 | 99 400 | 135 | **+79** | 51 | 0.998 |
| 2 | 98 616 | 529 | **+266** | 255 | 0.995 |
| 3 | 98 040 | 311 | **+30** | 265 | 0.993 |
| 4 | 97 465 | 287 | **+39** | 288 | 0.990 |
| 5 | 96 819 | 320 | **+9** | 326 | 0.986 |
| 6 | 96 165 | 322 | **+35** | 332 | 0.983 |
| 7 | 95 456 | 319 | **-1** | 390 | 0.979 |
| 8 | 94 638 | 378 | **+56** | 440 | 0.975 |
| 9 | 93 844 | 345 | **+26** | 449 | 0.970 |
| 10 | 92 962 | 342 | **-36** | 540 | 0.964 |
| 11 | 92 062 | 367 | **+22** | 533 | 0.959 |
| 12 | 91 096 | 376 | **+34** | 590 | 0.953 |
| 13 | 90 125 | 370 | **+3** | 601 | 0.947 |
| 14 | 89 096 | 420 | **+44** | 609 | 0.940 |
| 15 | 87 938 | 432 | **+62** | 726 | 0.933 |
| 16 | 86 724 | 425 | **+25** | 789 | 0.924 |
| 17 | 85 471 | 414 | 0 | 839 | 0.915 |
| 18 | 84 169 | 431 | **+21** | 871 | 0.906 |
| 19 | 82 818 | 368 | **+13** | 983 | 0.896 |
| 20 | 81 395 | 354 | 0 | 1069 | 0.884 |
| 21 | 79 866 | 366 | 0 | 1163 | 0.872 |
| 22 | 78 318 | 345 | 0 | 1203 | 0.859 |
| 23 | 76 710 | 361 | 0 | 1247 | 0.845 |
| 24 | 74 956 | 360 | 0 | 1394 | 0.830 |
| 25 | 73 055 | 400 | 0 | 1501 | 0.813 |
| 26 | 71 088 | 331 | 0 | 1636 | 0.795 |
| 27 | 68 989 | 326 | 0 | 1773 | 0.776 |
| 28 | 66 746 | 339 | 0 | 1904 | 0.755 |
| 29 | 64 276 | 312 | 0 | 2158 | 0.730 |
| 30 | 61 516 | 240 | 0 | 2520 | 0.702 |
| Total number of BCs / deaths |  | 10 872 | **+911** | 27 612 |  |

**Table S4:** Data on simulated cohort 4: 100,000 50-year-old women under U:S: life expectancy and SEER-age-specific BC risk over 30 years of virtual observation and endocrine prevention. The preventive therapy lasted two years. Therefore, newly initiated BCs are also eradicated immediately during therapy, leading to a prolonged BC reduction in the 16th and 17th years of follow-up.

| Year of diagnosis | Women  under  risk of BC | Age-specific  new BCs  per year  reduced by  prevented BCs  **(Cohort 4)** | **Difference of**  **BC cases compared to cohort 1** | **Competitive deaths according**  **to life expectancy ^1^** | **Expected**  **Survival** |
| --- | --- | --- | --- | --- | --- |
| 0 | 100 000 | 0 | 0 | 0 | 1 |
| 0.75 | 99 799 | 34 | **-29** | 167 | 0.999 |
| 1 | 99 723 | 25 | **-31** | 51 | 0.998 |
| 2 | 99 318 | 150 | **-113** | 255 | 0.995 |
| 3 | 98 922 | 131 | **-150** | 265 | 0.993 |
| 4 | 98 506 | 128 | **-120** | 288 | 0.990 |
| 5 | 98 027 | 153 | **-158** | 326 | 0.986 |
| 6 | 97 552 | 143 | **-144** | 332 | 0.983 |
| 7 | 97 002 | 160 | **-160** | 390 | 0.979 |
| 8 | 96 403 | 159 | **-163** | 440 | 0.975 |
| 9 | 95 795 | 159 | **-160** | 449 | 0.970 |
| 10 | 95 072 | 183 | **-195** | 540 | 0.964 |
| 11 | 94 360 | 179 | **-166** | 533 | 0.959 |
| 12 | 93 603 | 167 | **-175** | 590 | 0.953 |
| 13 | 92 832 | 170 | **-197** | 601 | 0.947 |
| 14 | 92 031 | 192 | **-184** | 609 | 0.940 |
| 15 | 91 110 | 195 | **-175** | 726 | 0.933 |
| 16 | 90 128 | 193 | **-207** | 789 | 0.924 |
| 17 | 89 079 | 210 | **-204** | 839 | 0.915 |
| 18 | 87 798 | 410 | 0 | 871 | 0.906 |
| 19 | 86 460 | 355 | 0 | 983 | 0.896 |
| 20 | 85 037 | 354 | 0 | 1069 | 0.884 |
| 21 | 83 508 | 366 | 0 | 1163 | 0.872 |
| 22 | 81 960 | 345 | 0 | 1203 | 0.859 |
| 23 | 80 352 | 361 | 0 | 1247 | 0.845 |
| 24 | 78 598 | 360 | 0 | 1394 | 0.830 |
| 25 | 76 697 | 400 | 0 | 1501 | 0.813 |
| 26 | 74 730 | 331 | 0 | 1636 | 0.795 |
| 27 | 72 631 | 326 | 0 | 1773 | 0.776 |
| 28 | 70 388 | 339 | 0 | 1904 | 0.755 |
| 29 | 67 918 | 312 | 0 | 2158 | 0.730 |
| 30 | 65 158 | 240 | 0 | 2520 | 0.702 |
| Total number of BCs / deaths |  | 7230 | **-2731** | 27 612 |  |

Table S5 Data sources and model assumptions

|  | Data Source | Measure/ derived measure | Assumptions |
| --- | --- | --- | --- |
| 1. | Chlebowski et.al. 2009 ^10^, Cuzick et al ^13^, Fisher et al. ^14^, Cuzick et al. ^15^, Weedon-Fekaer et al. ^12^, | - BCs can be described by their tumour diameter (TD). - Volume doubling time (VDT) is a central parameter for modeling tumour growth trajectories. - Volume doubling times (VDT) of 170 days in HR+ and of 72 days in HR- are deemed appropriate - 234 days is the 75%-percentile of VDT in tumours of a TD of 15 mm - A tumour with a diameter of 1 mm is composed of 1x10^6^ cells - A tumour with a diameter of 20 mm is composed of 8.6x10^9^ cells   $Growth duration in years\left( {TD}_{1}<{TD}_{2} \right)=\frac{log({TD}_{2}\div{TD}_{1})}{log(1.26)}\times\frac{VDT}{365}$ | - **1a)** A HR+ BC would take 15.3 years to grow from one cell with a diameter of 0.01 mm to a tumour of 20 mm at detection - This corresponds to 33 doublings - **1b)** A HR- BC of 20 mm TD would have a TD of 10 mm 0.6 years earlier, - A HR+ tumour of 20 mm of TD would have a TD of 10 mm 1.4 years earlier. - 15 years are a plausible simplified growth duration of a HR+ BC from initiation to detection |
| 2. | Cuzick et al. ^13^, Fisher et al. ^14^ | - HRT duration of 4 years - Growth acceleration factor of 2 corresponding to a VDT of 85 days - BC initiating risk factor of 2 (from Gail model) | Weighted mean of VDT of 150 days, considering the proportion of HR+ and HR- of 85 and 15% |
| 3. | Jemal et al. ^29^ U.S. SEER Cancer statistics review (1975-2002) | 1. 100 000 women aged 50 years are virtually observed over 30 years, BCs are randomly generated according to the age-specific incidence, life expectancy is considered. The age-specific incidence for the 50 years aged women is halved, to take screening mammography effects at the start into account 2. 100 000 women aged 50 years take HRT over 4 years and are observed over 30 years BCs and death are the same as in the control. The growth accelerating factor of 2 and the tumour development over 15 years are included in the modeling. 3. 100 000 women aged 50 years are under the same conditions as cohort 2. Additionally, the HRT-initiated BC risk factor of 2 is included in the modeling 4. 100 000 women aged 50 years take endocrine prevention over 2 years. A 50% treatment efficacy is modeled. | 1. Cohort 1: This group serves as a control without any intervention apart from further screening activities. 9961 BCs and 27 000 tumour-independent deaths occur within 30 years 2. Cohort 2: Due to growth acceleration the diagnosis of BCs (incidence) is shifted forward about two years within 19 years. This leads to an increased incidence up to the year 13 and decreased incidence after this up to the year 19. In the last decade of observation, the incidence is the same as in the control group. The total number of BCs is the same as in the control cohort 1. 3. Cohort 3: In the first 13 years of observation the BC incidence is the same as in cohort 2, with the growth acceleration and the diagnoses brought forward by 2 years. From year 14 on the newly initiated BCs with a development from first cell to the diagnosed tumour of 13-17 year (included 2 years of acceleration) increase the incidence compared to cohort 2. 4. Cohort 4: 2731 (27.3%) of prevalent BCs are eradicated prior to detection. The effect lasts for 17 years, after which the incidence is the same as in cohort 1 |
| 4. | Chlebowski et al. ^19^ Women’s Health Initiative Observational Study | The modeling above is done on four cohorts based on 8506 women, aged 63.6 years with HRT duration of 5.6 years. | See Figure 3 |
